# Tissue Eosinophil Counts as Predictors of Infliximab Response in Pediatric Inflammatory Bowel Disease: A Retrospective Nested Case Control Study

**DOI:** 10.1101/2025.09.08.25335331

**Authors:** Émile L’Heureux-Hubert, Samuel Sassine, Sophia Ferrante, Natacha Patey, Prévost Jantchou

**Affiliations:** Centre de recherche Azrieli du Centre Hospitalier Universitaire Sainte-Justine; Département de Médecine de l’Université de Montréal

## Abstract

**Background:** Infliximab is an effective therapy for pediatric inflammatory bowel disease (IBD), but a substantial proportion of patients experience primary non-response or relapse. Tissue eosinophilia has been implicated in treatment resistance, yet its predictive value in pediatric populations remains unclear.

**Methods:** We conducted a retrospective nested case-control study including 80 pediatric IBD patients treated with infliximab at CHU Sainte-Justine between 2014 and 2023 Relapse cases (n = 42) were defined by an increase in the short Pediatric Crohn’s Disease Activity Index (sPCDAI) ≥10, and compared with non-relapse controls (n = 38). Tissue eosinophil counts were quantified from diagnostic biopsies by blinded reviewers. Primary non-response was defined as a lack of clinical improvement before treatment change. Logistic regression and receiver operating characteristic (ROC) analyses were performed to assess associations between eosinophil counts and treatment outcomes.

**Results:** 80 patients were included in the study. Median age at diagnosis was 14.4 years (interquarile range [IQR] 3.65). Forty patients had a diagnosis of ulcerative colitis (UC, 50%) and the other half had a diagnosis of Crohn’s disease (CD). Baseline eosinophil counts were not associated with subsequent relapse (odds ratio [OR] 1.02, 95% CI 0.98-1.06, p = 0.31), and ROC analysis demonstrated poor discrimination (AUC 0.57). In contrast, higher eosinophil counts were associated with primary non-response in ulcerative colitis patients (OR 1.08, 95% CI 1.01-1.16, p = 0.03). A cutoff of a median of ≥ 36 eosinophils per high-power field among all segments yielded 85% sensitivity and 80% specificity for predicting primary non-response (AUC 0.82).

**Conclusion:** Tissue eosinophil counts do not predict relapse in pediatric IBD patients treated with infliximab. However, elevated counts may identify ulcerative colitis patients at risk of primary non-response, suggesting a role for histological stratification in guiding the selection of biologics. Prospective validation in larger cohorts is warranted.

## INTRODUCTION

Crohn’s disease (CD) and ulcerative colitis (UC) are the two most common forms of inflammatory bowel disease (IBD)(1). The incidence of Crohn’s disease in the Canadian pediatric population is among the highest in the world, at 13.9 per 100,000 children, compared with 2–5 per 100,000 children in other regions(2, 3). Moreover, this incidence has been steadily increasing over time(4).

Pediatric inflammatory bowel disease (IBD) is a chronic, relapsing condition that imposes a substantial burden on patients and health systems (5). The incidence of both Crohn’s disease (CD) and ulcerative colitis (UC) has been rising worldwide, with increasing recognition in children and adolescents (6). Early and effective treatment is essential to control inflammation, prevent complications, and optimize growth and quality of life. Among available biologics, infliximab, an anti–tumour necrosis factor alpha (TNF-α) monoclonal antibody, is a cornerstone therapy for moderate to severe pediatric IBD (6). Despite its proven efficacy, up to one-third of patients experience primary non-response during induction, and nearly half relapse during maintenance, highlighting the urgent need for reliable predictive biomarkers (7).

Primary non-response and secondary loss of response to infliximab remain major clinical challenges in pediatric IBD. Reported rates vary, but approximately 20–30% of children fail to achieve clinical remission during induction, and 30–50% relapse within two years of maintenance therapy(8). Pharmacokinetic factors are key in prospective pediatric study, children with week-14 infliximab through levels <5 µg/mL had significantly higher chances of relapse within the first year compared with those with higher concentrations (9, 10). Immunogenicity is also important, with anti-drug antibodies detected in up to 20–40% of patients, correlating with both infusion reactions and treatment failure (11). Clinical predictors such as perianal disease in Crohn’s (associated with nearly double the risk of loss of response), extensive colitis in UC, and elevated baseline CRP have all been linked to poorer outcomes (12, 13). In a multicenter cohort observed that 32% of pediatric patients treated with anti-TNF agents experienced primary non-response (8%) or early relapse (24% in the first three years), despite optimized treatment, highlighting the urgent need for predictive biomarkers(14). In pediatric luminal CD, Sassine et al. observed that elevated mucosal eosinophilia was associated with clinical relapse (aHR = 1.36, *P* = 0.02) (15).

Eosinophils are multifunctional immune cells implicated in type 2-driven inflammation and tissue remodeling. Although traditionally associated with allergic disorders, eosinophils are increasingly recognized as contributors to the pathogenesis of IBD(16, 17). They accumulate in inflamed mucosa, release cytotoxic granule proteins, and interact with other immune cells, including T helper 2 (Th2) lymphocytes and mast cells (18, 19). Several studies have suggested that mucosal eosinophilia correlates with disease activity, particularly in UC, and may influence therapeutic response (19, 20). In adults, high eosinophil counts have been linked to treatment failure with vedolizumab and other biologics (21, 22). However, the evidence remains inconsistent, and data in children are scarce (18, 23).

Identifying histological predictors of infliximab response in pediatric IBD could improve patient stratification and guide treatment decisions. Biopsy analysis is routinely performed at diagnosis, providing an opportunity to integrate histopathology into clinical algorithms without additional burden. Yet, the predictive role of tissue eosinophil counts for infliximab outcomes in the pediatric setting has not been systematically evaluated. (24)

The present study aimed to assess whether tissue eosinophil counts at diagnosis are associated with infliximab outcomes in a pediatric IBD cohort, with the primary objective of evaluating whether tissue eosinophil counts predict primary non-response to infliximab during the induction phase, and the secondary objective of investigating whether tissue eosinophil counts can guide therapeutic decision-making for achieving durable remission in pediatric IBD.

## METHODS

### Study design and population

We conducted a retrospective nested case–control study within the pediatric IBD cohort at CHU Sainte-Justine (Montreal, Canada). Patients diagnosed with colonic CD or UC between 2014 and 2023 were eligible if they received infliximab therapy and had diagnostic endoscopic biopsies available for review. Patients with indeterminate colitis or missing follow-up data were excluded. The study was approved by the institutional research ethics board.

### Outcomes

Cases were children who initiated infliximab treatment within 30 days of diagnosis and experienced a clinical relapse, defined as an increase of ≥10 points in sPCDAI requiring either a change in biological therapy or the addition of corticosteroid treatment within the first year after diagnosis.

Controls were children who initiated infliximab treatment within 30 days of diagnosis and maintained remission during the first year, without meeting the relapse criteria.

### Histological assessment

Formalin-fixed paraffin-embedded biopsies obtained at diagnosis were reviewed. A pathologist and a trained investigator, blinded to clinical outcomes, quantified eosinophil counts as the peak number of eosinophils per high-power field (HPF, 400× magnification) for each segment (ileon, ascending colon, transverse colon, descending colon, sigmoid and rectum). Inter-rater reliability was excellent (intraclass correlation coefficient = 0.94) The eosinophil count per HPF was compared for all segments individually between responders and relapsing participants before using the median of all segments for further analysis.

### Treatment protocol

Infliximab was administered intravenously at a standard induction regimen (5 to 10 mg/kg at weeks 0, 2, and 6), in the first 30 days following diagnosis followed by maintenance infusions every 8 weeks. Dose escalation or interval shortening was permitted at the treating physician’s discretion.

### Data collection

Demographic and clinical data were extracted from electronic medical records, including age at diagnosis, sex, IBD subtype (Crohn’s disease or ulcerative colitis), disease location and behaviour according to the Paris classification, and prior treatment history including corticosteroids and other medications.

Potential confounding factors considered in the analysis included baseline disease severity (CRP), prior corticosteroids exposure, age, and IBD subtype all collected.

### Statistical analysis

The primary objective to evaluate whether tissue eosinophil counts predict primary non-response to infliximab during induction will be assessed using logistic regression. In this model, primary non-response (yes/no) is the dependent variable, and tissue eosinophil count (continuous or categorized) is the main independent variable. The analysis will be adjusted for potential confounding factors, such as age, sex, baseline disease severity (CRP), history of corticosteroids use and infliximab dose at induction. Results will be reported as odds ratios (ORs) with 95% confidence intervals (CIs).

The secondary objective to investigate whether tissue eosinophil counts predict relapse during maintenance therapy will similarly be analyzed using logistic regression, with relapse (yes/no) as the outcome. Cases and controls are defined as described previously. Adjustment for the same set of confounders will be performed to account for factors that could influence both eosinophil counts and risk of relapse. Results will be reported as odds ratios (ORs) with 95% confidence intervals (CIs).

For exploratory analyses correlation analyses will be used to examine the relationship between tissue eosinophil counts and circulating eosinophil levels.

Additional analyses include receiver operating characteristic (ROC) curve analysis to evaluate the discriminative ability of tissue eosinophil counts for predicting primary non-response and relapse, and to identify optimal cutoff values. Baseline characteristics will be compared using chi-square or Fisher’s exact test for categorical variables, and Student’s t-test or Wilcoxon rank-sum test for continuous variables, as appropriate. Statistical significance is defined as p < 0.05, and all analyses will be performed using SAS version 9.4.

## RESULTS

### Cohort characteristics

Eighty pediatric patients with IBD treated with infliximab were included, comprising 40 with Crohns disease (50%) and 40 with ulcerative colitis (50%). Median age at diagnosis was 14.4 years (IQR 3.65), and 70% were male. Forty-two patients remained in remission, and 38 experienced relapses (among these 38, five patients had a primary non-response). Baseline demographic and clinical characteristics were similar between relapse and non-relapse groups (Table 1).

**TABLE 1.** Distribution of demographic, clinical and histological variables in patients who maintained infliximab response.

|  | Sustained response to infliximab N=42* | Relapse on infliximab requiring a change in treatment N=38* |
| --- | --- | --- |
| Demographics & Baseline clinical characteristics |  |  |
| Male sex, n (%) | 19 (45.2) | 20 (52.6) |
| IBD type: Ulcerative colitis, n (%) | 20 (47.6) | 20 (52.6) |
| Age at diagnosis, years (median [IQR]) | 14.4 [3.9] | 14.5 [3.6] |
| BMI z-score (median [IQR]) | -0.7 [2.1] | -0.6 [2.3] |
| Height z-score (median [IQR]) | -0.1 [1.6] | -0.1 [1.1] |
| Weight z-score (median [IQR]) | -0.5 [2.2] | -0.4 [1.5] |
| Crohn's disease location (Paris classification) |  |  |
| Colonic (L2), n (%) | 5 (22.7) | 6 (33.3) |
| Ileocolonic (L3), n (%) | 17 (77.3) | 12 (66.7) |
| Localisation of Ulcerative Colitis (Paris classification) |  |  |
| Proctitis (E1), n (%) | 1 (5) | 0 (0) |
| Left-sided colitis (E2), n (%) | 1 (5) | 2 (10) |
| Extensive colitis, distal to cecum (E3), n (%) | 0 (0) | 2 (10) |
| Pancolitis (E4), n (%) | 18 (90) | 16 (80) |
| UC severity at diagnosis (S0), n (%) | 11 (55) | 8 (40) |
| Extraintestinal manifestations, n (%) | 11 (52.4) | 8 (44.4) |
| PUCAI score at diagnosis (median [IQR]) | 60.0 [17.5] | 67.5 [20.0] |
| PCDAI score at diagnosis (median [IQR]) | 47.5 [15.0] | 50.0 [11.3] |
| SES-CD score at diagnosis (median [IQR]) | 20.0 [12.0] | 13.5 [12.0] |
| Histological characteristics |  |  |
| Architectural distortion present, n (%) | 31 (73.8) | 35 (92.1) |
| Lymphoid follicles present, n (%) | 11 (26.2) | 12 (31.6) |
| Granulomas present, n (%) | 4 (9.5) | 4 (10.5) |
| Median eosinophil count (all sites) (median [IQR]) | 21.8 [14.3] | 22.5 [19.6] |
| Eosinophil count, ileum (median [IQR]) | 17.5 [24.5] | 20.5 [17.0] |
| Eosinophil count, ascending colon (median [IQR]) | 20.0 [25.5] | 22.0 [20.0] |
| Eosinophil count, transverse colon (median [IQR]) | 20.3 [21.5] | 20.3 [21.2] |
| Eosinophil count, descending colon (median [IQR]) | 19.0 [17.0] | 24.8 [31.0] |
| Eosinophil count, sigmoid colon (median [IQR]) | 16.5 [17.5] | 22.5 [23.5] |
| Eosinophil count, rectum (median [IQR]) | 14.3 [23.0] | 16.0 [25.5] |
| Variables associated to infliximab treatment |  |  |
| Delay before induction (days) (median [IQR]) | −4.0 [6.0] | −3.5 [10.0] |
| Induction IFX dose (mg/kg) (median [IQR]) | 9.6 [1.9] | 9.4 [4.1] |
| IFX trough level during inductions (µg/mL) (median [IQR]) | 16.4 [17.4] | 24.6 [22.8] |
| Median Maintenance IFX dose (mg/kg) (median [IQR]) | 10.4 [2.5] | 10.0 [2.9] |
| Median IFX trough level during maintenance (µg/mL) (median [IQR]) | 15.6 [11.1] | 14.8 [13.9] |
| Median anti-IFX antibody titer (AU/mL) (median [IQR]) | 6.0 [15.5] | 6.8 [23.5] |
| Anti-IFX antibodies >10 AU/mL, n (%) | 5 (12.5) | 6 (15.8) |
| Low induction through level, n (%)** | 8 (20) | 6 (15.8) |
| Low maintenance trough level, n (%)*** | 4 (10) | 6 (15.8) |
| Prior Systemic corticosteroid use, n (%) | 36 (85.7) | 32 (84.2) |

\* All percentages are columns proportions \*\* IFX through levels under 25, 15 or 10 for either the 0, 2 and 6 doses respectively. \*\*\* IFX trough level under 5 BMI : Body mass index, IQR : interquartile range , PUCAI: Pediatric Ulcerative Colitis Activity Index, PCDAI: Pediatric Crohn’s Disease Activity Index, UC: Ulcerative Colitis SES-CD: Simple Endoscopic Score – Crohn’s Disease , IFX, Infliximab

Regarding relapse and outcomes, among the 80 included patients, 5 (13□%) experienced primary non-response to infliximab. Among the 38 patients who relapsed and required a treatment change, 16 (42.1□%) received corticosteroids, 11 (28.9□%) were switched to ustekinumab, 7 (18.4□%) to vedolizumab, and 4 (10.5□%) to tofacitinib. Clinical activity at the time of relapse was moderate to high, with a median PUCAI score of 50.0 [IQR 30.0] in patients with ulcerative colitis, a median PCDAI score of 35.0 [IQR 27.5] in patients with Crohn’s disease, and a median SES-CD score of 11.0 [IQR 15.0], reflecting persistent intestinal inflammation at relapse.

A Pearson correlation analysis was performed to evaluate the association between tissue eosinophil counts and blood eosinophil counts. A weak but statistically significant positive correlation was observed (r = 0.275, p = 0.0141).

Patients with ulcerative colitis had significantly higher overall eosinophil counts compared to those with Crohn’s disease (median 23.6 vs 15.5, *p*=0.0068). This difference was consistent across most colonic segments, including the ascending, transverse, descending, and sigmoid colon, as well as the rectum, where the largest gap was observed (29 vs 9, *p*=0.0005). In contrast, the ileum showed the opposite pattern, with higher eosinophil counts in Crohn’s disease (23 vs 16.5, *p*=0.0364) (Table 2).

**TABLE 2.** Comparison of eosinophil counts by intestinal segment in UC and CD patients.

|  | Patients with UC N=40 | Patients with CD N=40 | P-value |
| --- | --- | --- | --- |
| Median eosinophil count (all sites) (median [IQR]) | 23.6 [19.4] | 15.5 [14.0] | 0.0068 |
| Eosinophil count, ileum (median [IQR]) | 16.5 [15.0] | 23.0 [23.0] | 0.0364 |
| Eosinophil count, ascending colon (median [IQR]) | 23.5 [22.5] | 18.5 [23.5] | 0.0469 |
| Eosinophil count, transverse colon (median [IQR]) | 26.0 [19.5] | 15.5 [14.0] | 0.0085 |
| Eosinophil count, descending colon (median [IQR]) | 25.0 [29.5] | 15.5 [21.5] | 0.0309 |
| Eosinophil count, sigmoid colon (median [IQR]) | 26.5 [25.5] | 14.0 [16.0] | 0.0079 |
| Eosinophil count, rectum (median [IQR]) | 29.0 [21.5] | 9.0 [10.5] | 0.0005 |

IQR : interquartile range UC : ulcerative colitis CD : Crohn’s disease

### Relapse

Baseline tissue eosinophil counts did not differ significantly between patients who relapsed and those who did not (median 22.5 vs 21.75 eosinophils/HPF, p = 0.21). No difference was detected in the number of eosinophils between the two groups, even when stratifying the analyses for each segment individually. In logistic regression, eosinophil counts were not associated with relapse (OR 1.02, 95% CI 0.98; 1.05, p = 0.3083) (Table 3). ROC analysis confirmed poor discriminatory ability (AUC 0.57; 95% CI 0.44; 0.71).

**TABLE 3.** Binary logistic regression for the variables independently associated with a relapse to infliximab treatment necessitating the addition of a new treatment.

|  | Univariate Odds Ratio |  | Adjusted Odds Ratio* |  |
| --- | --- | --- | --- | --- |
|  | OR (95% CI) | P-Value | OR (95% CI) | P-Value |
| Age at diagnosis, years | 1.02 (0.92 ; 1.12) | 0.7817 | 0.96 (0.78 ; 1.18) | 0.6946 |
| Sex (female vs male) | 0.74 (0.31 ; 1.79) | 0.5092 | 0.87 (0.33 ; 2.26) | 0.7727 |
| IBD type (UC vs CD) | 1.22 (0.51 ; 2.94) | 0.6545 | 1.83 (0.59 ; 5.65) | 0.2957 |
| Median eosinophil count (all sites) | 0.87 (0.74 ; 1.03) | 0.2078 | 1.02 (0.98; 1.05) | 0.3083 |
|  | OR (95% CI) | P-Value | OR (95% CI) | P-Value |
| Induction dose of infliximab mg/kg | 0.74 (0.55 ; 0.99) | 0.0350 | 0.76 (0.58 ; 0.99) | 0.0386 |
| CRP at diagnosis | 1 (0.99;1.01) | 0.4484 | 1 (0.99 ; 1.01) | 0.6456 |
| Exposure to corticosteroids | 1.24 (0.36 ; 4.31) | 0.7301 | 0.83 (0.19 ; 3.64) | 0.8053 |

OR : odds ratio ORa : adjusted odds ratio UC : ulcerative colitis CD : Crohn's disease CRP : C-reactive protein
\* Results from a multivariate linear logistic regression model adjusted for all variables included in the univariate analyses.

**TABLE 4.** Binary logistic regression for the variables independently associated with a primary non-response to infliximab in CU patients.

|  | Univariate Odds Ratio |  | Adjusted Odds Ratio* |  |
| --- | --- | --- | --- | --- |
|  | OR (95% CI) | P-Value | ORa (95% CI) | P-Value |
| Age at diagnosis, years | 1.20 (0.78 ; 1.87)) | 0.4075 | 1.09 (0.59 ; 1.99) | 0.7883 |
| Sex (female vs male) | 0.22 (0.02 ; 2.05) | 0.1831 | 0.27 (0.01 ; 5.38) | 0.3932 |
| Median eosinophil count (all sites) | 1.08 (1.02 ; 1.14) | 0.0089 | 1.13 (1.01 ; 1.27) | 0.0342 |
| Induction dose of infliximab mg/kg | 0.99 (0.62 ; 1.57) | 0.1855 | 0.61 (0.28 ; 1.33) | 0.2118 |
| CRP at diagnosis | 1.01 (1.00 ; 1.03) | 0.1339 | 1.03 (0.99 ; 1.07) | 0.1154 |
| Exposure to corticosteroids** | . | . | . | . |

OR : odds ratio ORa : adjusted odds ratio CRP : C-reactive protein
\* Results from a multivariate logistic regression model adjusted for all variables included in the univariate analyses.
\*\* All patients included in this analysis had prior corticosteroid exposure

### Primary non-response

Of the 80 patients, 5 (6.25%) met criteria for primary non-response. These patients had significantly higher median baseline eosinophil counts compared with responders (40.3 vs 21.8 eosinophils/HPF, p = 0.0139) or non-responder (40.3 vs 21.8, p=0.0323). Circulating eosinophiles were not higher in primary non-responders (medians 0.3 vs 0.9, p=0.16).

In multivariable logistic regression adjusted for age, sex, and induction dose of infliximab, eosinophil counts were independently associated with primary non-response (OR 1.13 per eosinophil/HPF, 95% CI 1.01; 1.27, p = 0.0342).

ROC analysis identified an optimal cutoff of ≥36 eosinophils/HPF using the highest Youden index, yielding 85% sensitivity and 80% specificity (AUC 0.82, 95% CI 0.66; 0.97) (Figure 1).

**FIGURE 1.**
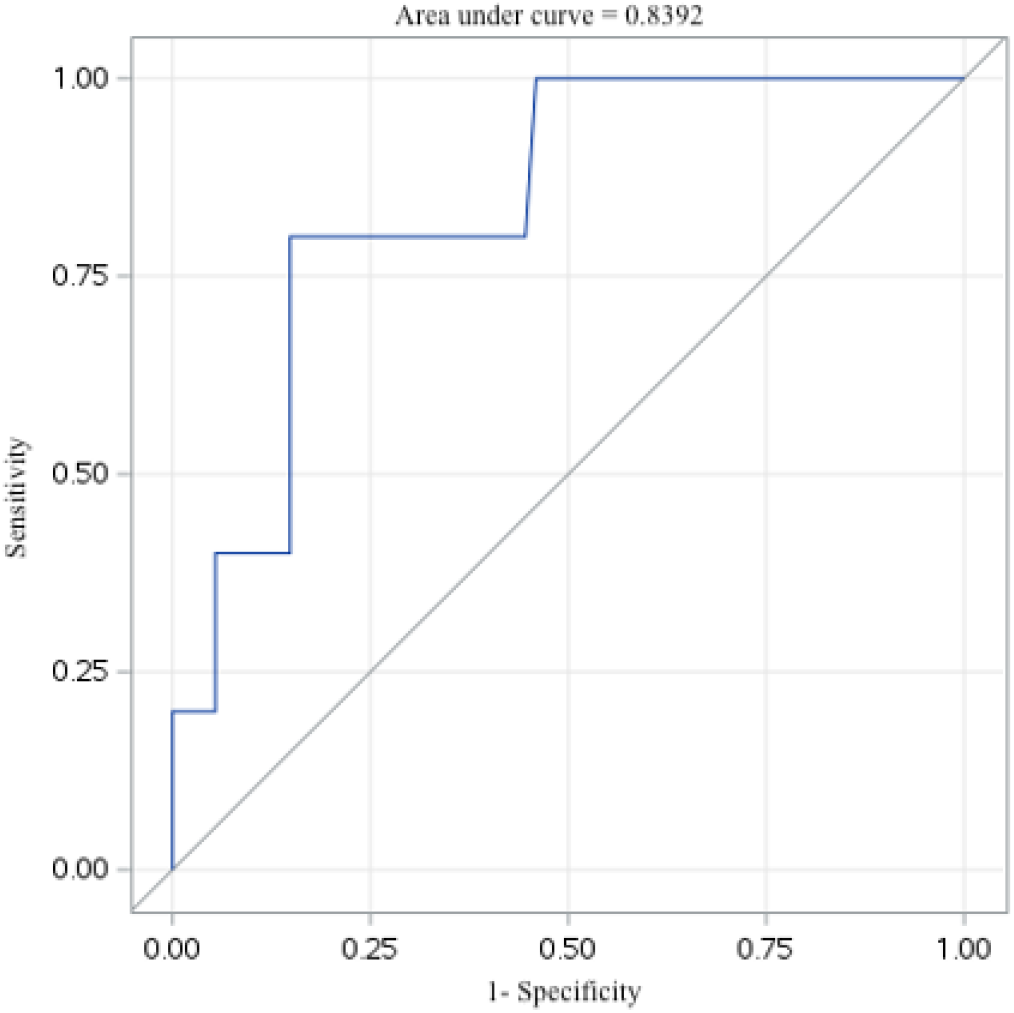
Receiver operating characteristic (ROC) curve for baseline eosinophils counts predicting primary non-response in ulcerative colitis.

## DISCUSSION

In this retrospective cohort of pediatric IBD patients treated with infliximab, we found that tissue eosinophil counts at diagnosis were not associated with relapse during maintenance therapy. However, higher eosinophil counts were significantly associated with primary non-response in ulcerative colitis, with an optimal cutoff of ≥36 eosinophils per high-power field providing good sensitivity and specificity. These findings suggest that mucosal eosinophilia may contribute to early treatment resistance in UC, while mechanisms underlying relapse appear to be independent of eosinophil activity.

Our results align with the emerging view that eosinophils play a distinct role in the immunopathology of UC compared with Crohn’s disease (25). Eosinophils accumulate preferentially in the colonic mucosa, where they release pro-inflammatory mediators such as major basic protein, eosinophil peroxidase, and cytokines including IL-5 and IL-13 (26). These factors promote epithelial barrier disruption, fibrosis, and amplification of type 2 inflammation (25, 27). It is plausible that such pathways reduce the effectiveness of TNF-α blockade, explaining the observed association with primary non-response (22). In contrast, relapse during infliximab maintenance is often driven by immunogenicity, low through levels, or activation of TNF-independent pathways, which may account for the lack of predictive value in this setting (28).

Several adult studies support the role of eosinophils in biological resistance. High mucosal eosinophil counts have been linked to non-response to vedolizumab in UC (21, 22), and peripheral eosinophilia has been associated with adverse outcomes in anti-TNF–treated patients (29). However, data on children are limited(23). Our findings contribute novel evidence from a pediatric cohort, highlighting the potential of tissue eosinophils as predictive biomarkers at treatment initiation. Importantly, the effect was observed specifically in UC, underscoring disease-specific mechanisms that merit further study.

From a clinical perspective, eosinophil quantification is a simple, inexpensive, and widely available histological measure (30). If validated, it could be incorporated into diagnostic pathology reports to help guide therapeutic decision-making. For instance, pediatric UC patients with high eosinophil counts at diagnosis may benefit from closer monitoring of early infliximab response, or consideration of alternative biologics such as ustekinumab or vedolizumab as new evidence points towards a role of eosinophils as a biomarker for those monitoring response to these treatments in IBD(22, 31). Integrating histological markers with clinical and pharmacological data could ultimately improve individualized treatment strategies.

This study has some limitations. Its retrospective design and modest sample size limit generalizability and results should be interpreted with caution. Although inter-rater reliability for eosinophil counts was excellent (ICC), histological variability remains inherent to biopsy sampling. We did not assess serum or fecal biomarkers, such as eotaxin or calprotectin, which may complement tissue eosinophilia in predictive models. Additionally, our ability to analyze subgroups was limited, particularly for Crohn’s disease, as none of our primary nonresponders had Crohn’s disease. Future studies should validate these findings prospectively in larger, multicenter pediatric cohorts. Integrating eosinophil counts with molecular biomarkers, pharmacokinetics, and genetic factors may yield more robust predictive algorithms. Mechanistic studies exploring the interaction between eosinophil-driven pathways and TNF blockade could also clarify therapeutic resistance (16, 32). Ultimately, such research may enable precision medicine approaches, ensuring that children with IBD receive the most effective therapy from the outset.

## CONCLUSION

In this pediatric IBD cohort, tissue eosinophil counts at diagnosis were not predictive of relapse during infliximab maintenance. However, higher eosinophil counts were significantly associated with primary non-response in ulcerative colitis, suggesting a role for mucosal eosinophilia in early treatment resistance. If validated, biopsy-based eosinophil quantification could serve as a simple histological biomarker to guide therapeutic stratification in pediatric UC. Prospective multicenter studies are warranted to confirm these findings and explore integration with molecular and pharmacological predictors.

## Data Availability

All data produced in the present study are available upon reasonable request to the authors

## Supplementary Data

**TABLE 5.**
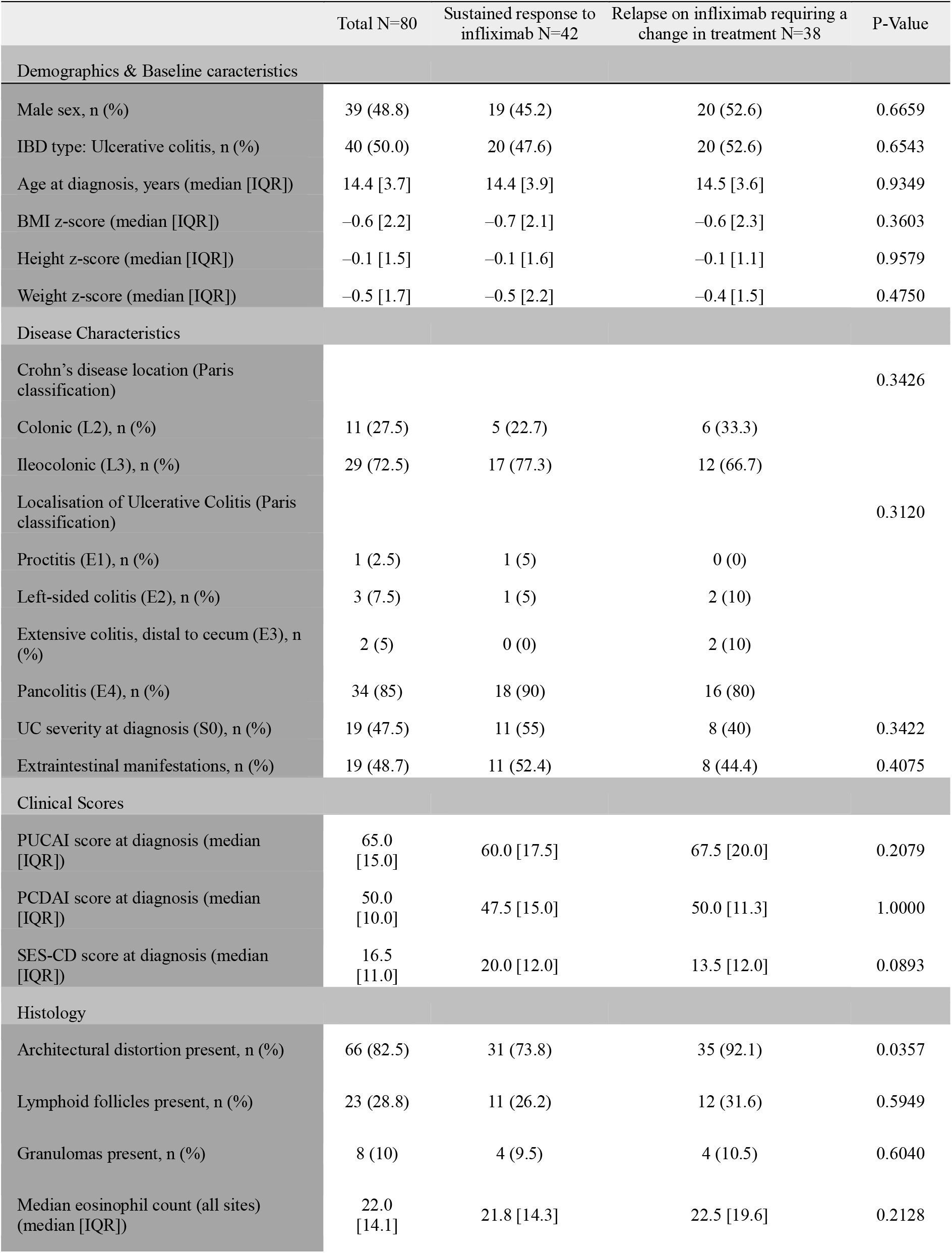
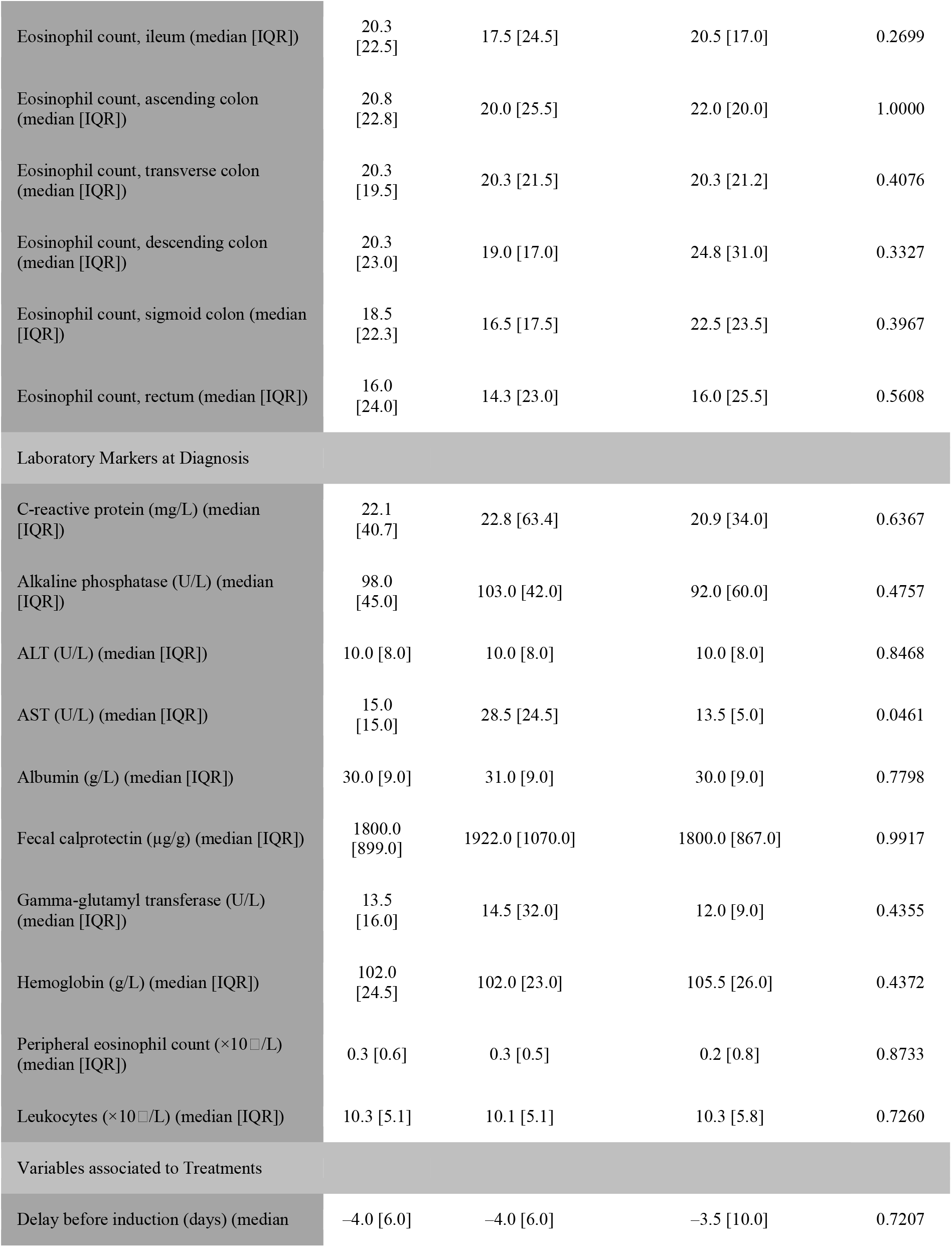
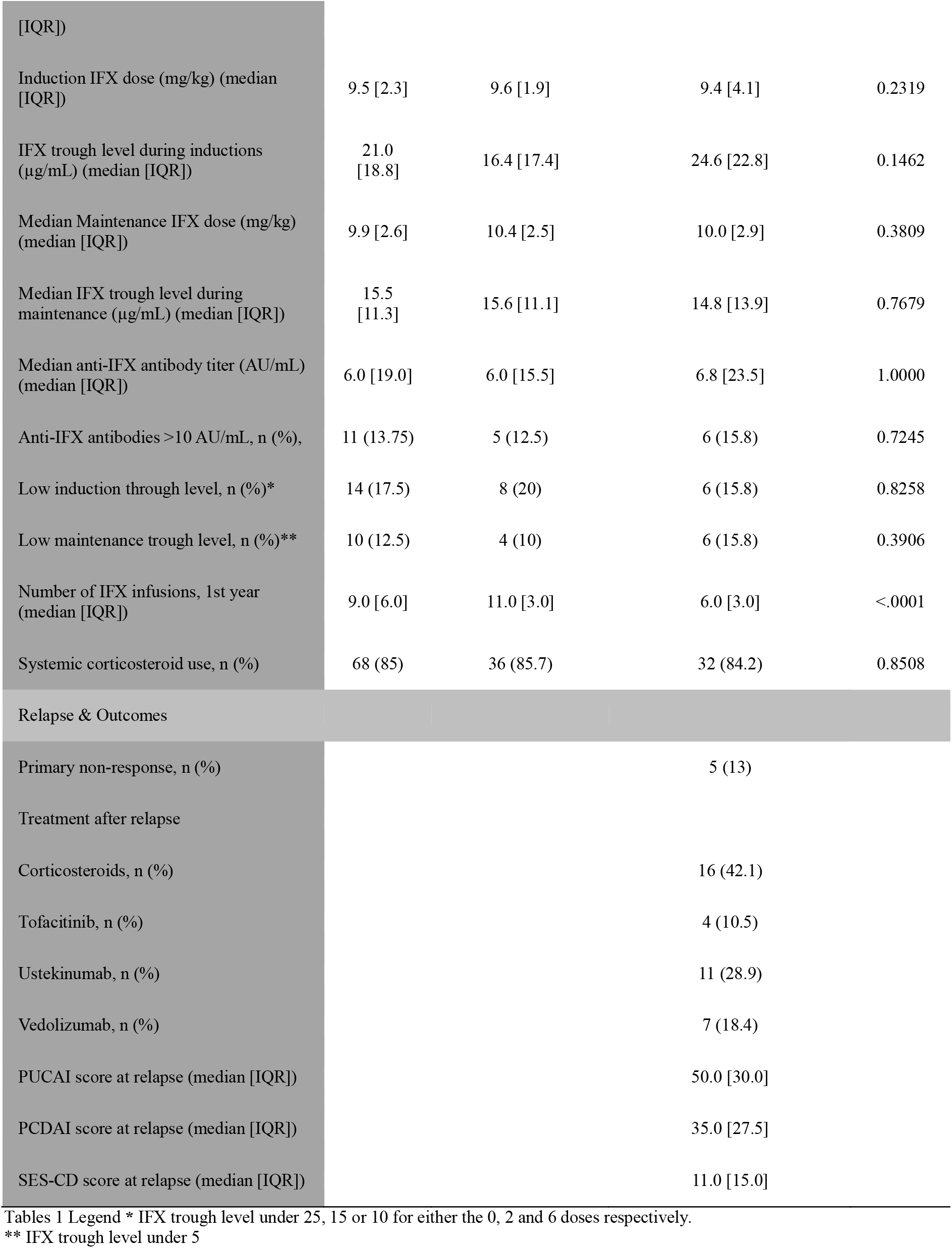
Distribution of demographic, clinical and histological variables in patients who maintained infliximab response.

